# Self-Reported Mask Use among Persons with or without SARS CoV-2 Vaccination —United States, December 2020–August 2021

**DOI:** 10.1101/2022.04.06.22273448

**Authors:** Lydia E. Calamari, Ashley H. Tjaden, Sharon L. Edelstein, William S. Weintraub, Roberto Santos, Michael Gibbs, Johnathan Ward, Michele Santacatterina, Alain G. Bertoni, Lori M. Ward, Sharon Saydah, Ian D. Plumb, Michael S. Runyon, COVID-19 Community Research Partnership Study Group

**Affiliations:** Atrium Health, Charlotte, NC; The Biostatistics Center, Milken Institute School of Public Health, George Washington University, Washington, DC; MedStar Health Research Institute and Georgetown University, Washington, DC; University of Mississippi Medical Center, Jackson, MS; Centers for Disease Control and Prevention, Atlanta, GA; Vysnova Partners, Inc.; Wake Forest School of Medicine, Winston-Salem, NC

**Keywords:** COVID-19, mask use, vaccination

## Abstract

Wearing a facemask can help to decrease the transmission of COVID-19. We investigated self-reported mask use among subjects aged 18 years and older participating in the COVID-19 Community Research Partnership (CRP), a prospective longitudinal COVID-19 surveillance study in the mid-Atlantic and southeastern United States. We included those participants who completed ≥5 daily surveys each month from December 1, 2020 through August 31, 2021. Mask use was defined as self-reported use of a face mask or face covering on every interaction with others outside the household within a distance of less than 6 feet. Participants were considered vaccinated if they reported receiving ≥1 COVID-19 vaccine dose. Participants (n=17,522) were 91% non-Hispanic White, 68% female, median age 57 years, 26% healthcare workers, with 95% self-reported receiving ≥1 COVID-19 vaccine dose through August; mean daily survey response was 85%. Mask use was higher among vaccinated than unvaccinated participants across the study period, regardless of the month of the first dose. Mask use remained relatively stable from December 2020 through April (range 71–80% unvaccinated; 86–93% vaccinated) and declined in both groups beginning in mid-May 2021 to 34% and 42% respectively in June 2021; mask use has increased again since July 2021. Mask use by all was lower during weekends and on Christmas and Easter, regardless of vaccination status. Independent predictors of higher mask use were vaccination, age ≥65 years, female sex, racial or ethnic minority group, and healthcare worker occupation, whereas a history of self-reported prior COVID-19 illness was associated with lower use.

## Introduction

Mask use is a primary public health method to help prevent the spread of SARS-CoV-2, especially when in close contact with individuals outside of the household.^1^ Systematic reviews and meta-analyses support mask use among other strategies to reduce the incidence of SARS-CoV-2 infection and ultimately COVID-19 mortality.^2^ Furthermore, many studies have supported the role of mask use by mask mandate to decrease both infection with SARS-CoV-2and the rate of hospitalization in patients with COVID-19.^3^ Vaccination against COVID-19 became available in the United States in December 2020 and more widely over the subsequent months. However, there is limited information on rates of mask use over time or by vaccination status. Using prospectively-collected data from the COVID-19 Community Research Partnership (CRP), we report changes in mask use over time among vaccinated and unvaccinated participants.

## Methods

The CRP is a prospective, multi-site cohort syndromic COVID-19 surveillance study of a convenience sample of participants enrolled primarily through direct email outreach at ten healthcare systems from the mid-Atlantic and southeastern United States, April 2020 through June 2021 (http://www.covid19communitystudy.org/). Informed consent, baseline demographic information and healthcare worker status were obtained. Daily online email or text surveys asked about COVID-19 exposures, symptoms, and mask use. Mask use was reported as “Yes”, “No” or “No interactions” to the question: “In the last 24 hours, have you worn a face mask or face covering every time you interacted with others (not in your household) within a distance of less than 6 feet?” We included participants 18 years and older, enrolled by December 2020 (when COVID-19 vaccination first became available in the United States), who completed daily surveys ≥5 times each month, December 2020 through August 2021. For the purpose of this analysis, participants were considered vaccinated if they reported receiving at least one dose of any COVID-19 vaccine by August 31, 2021. This study was reviewed and approved by the Wake Forest Institutional Review Board (IRB), which served as the central IRB for this study (See 45 C.F.R. part 46; 21 C.F.R. part 56). The study is registered with ClinicalTrials.gov, NCT04342884.

### Statistical Methods

Participants were categorized into month of self-reported receipt of the first dose of a COVID-19 vaccine, or lack of self-reported COVID-19 vaccination. Mask use was defined as “yes” to the mask use question; responses of “no interactions” were excluded from the main analysis but are included in supplemental material. The rate of mask use was calculated as the proportion of daily responses per day and per week. Multivariable logistic regression models were used to assess whether vaccination status was associated with the answer of “yes” to the mask question in the first weeks of March, June, and August 2021 to assess the relationship at various times during the pandemic. Models were adjusted for self-reported prior COVID-19 illness, age group, sex, race/ethnicity, and healthcare worker occupation. Analyses were performed using R (V.4.0.3).

## Results

Of 36,220 adult CRP volunteers enrolled by December 2020, 17,522 (48%) met the survey completion criteria (Supplemental Table 2 and Supplemental Figure 1). The daily survey response rate was 85% (Supplemental Figure 2) and 16,707 (95.3%) reported receiving ≥1 COVID-19 vaccine dose. Most participants were non-Hispanic White (90.7%) and female (68.1%). Median age was 57 years and 25.6% were healthcare workers. Older participants and males were more likely to be vaccinated (Table 1). Among vaccinated participants, 47.7% (n=7970) reported their first dose in December 2020 or January 2021, 43.4% (n=7257) in February or March 2021, 6.4% (n=1074) in April 2021 and <1% per month in May through August 2021. Only 4.7% (n=815) did not report vaccination.

**Table 1:**
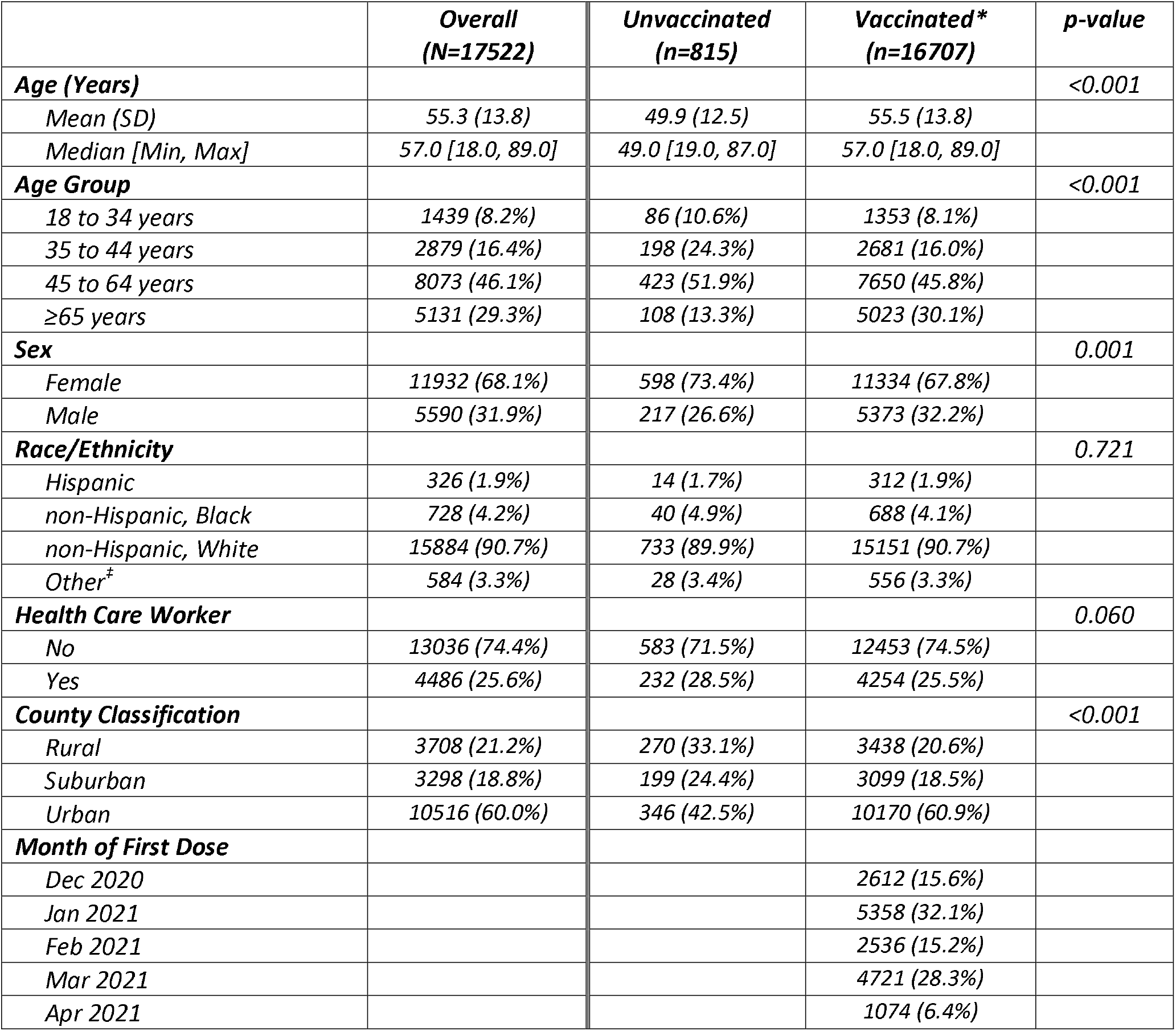
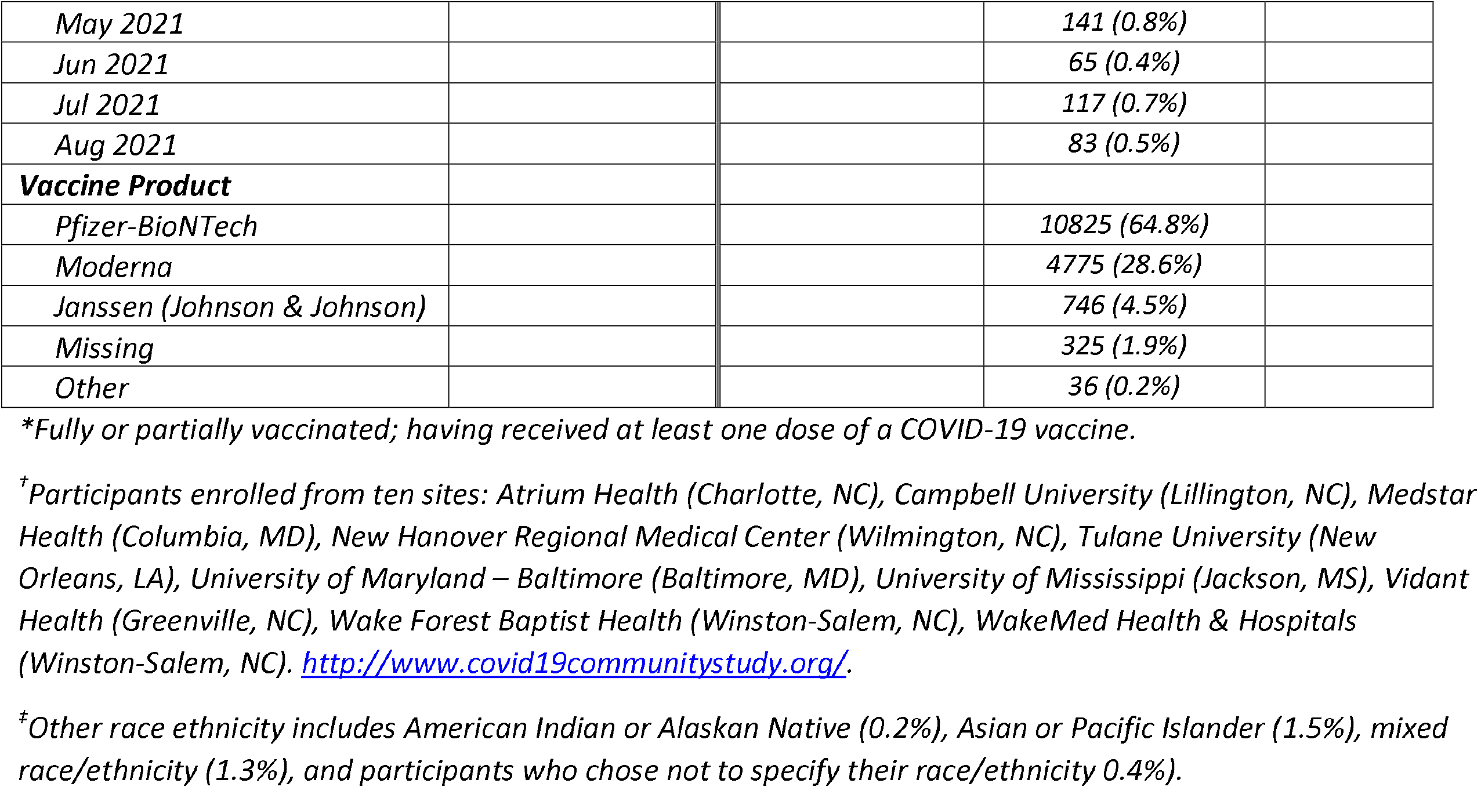
Distribution of study participants by vaccination status* as of August 31, 2021, COVID-19 Community Research Partnership^†^

|  | **Overall**<br>**(N=17522)** | **Unvaccinated**<br>**(n=815)** | **Vaccinated\***<br>**(n=16707)** | **p-value** |
| --- | --- | --- | --- | --- |
| **Age (Years)** |  |  |  | **<0.001** |
| Mean (SD) | 55.3 (13.8) | 49.9 (12.5) | 55.5 (13.8) |  |
| Median [Min, Max] | 57.0 [18.0, 89.0] | 49.0 [19.0, 87.0] | 57.0 [18.0, 89.0] |  |
| **Age Group** |  |  |  | **<0.001** |
| 18 to 34 years | 1439 (8.2%) | 86 (10.6%) | 1353 (8.1%) |  |
| 35 to 44 years | 2879 (16.4%) | 198 (24.3%) | 2681 (16.0%) |  |
| 45 to 64 years | 8073 (46.1%) | 423 (51.9%) | 7650 (45.8%) |  |
| $\geq 65$ years | 5131 (29.3%) | 108 (13.3%) | 5023 (30.1%) | |
| **Sex** |  |  |  | **0.001** |
| Female | 11932 (68.1%) | 598 (73.4%) | 11334 (67.8%) |  |
| Male | 5590 (31.9%) | 217 (26.6%) | 5373 (32.2%) |  |
| **Race/Ethnicity** |  |  |  | **0.721** |
| Hispanic | 326 (1.9%) | 14 (1.7%) | 312 (1.9%) |  |
| non-Hispanic, Black | 728 (4.2%) | 40 (4.9%) | 688 (4.1%) |  |
| non-Hispanic, White | 15884 (90.7%) | 733 (89.9%) | 15151 (90.7%) |  |
| Other<sup>‡</sup> | 584 (3.3%) | 28 (3.4%) | 556 (3.3%) |  |
| **Health Care Worker** |  |  |  | **0.060** |
| No | 13036 (74.4%) | 583 (71.5%) | 12453 (74.5%) |  |
| Yes | 4486 (25.6%) | 232 (28.5%) | 4254 (25.5%) |  |
| **County Classification** |  |  |  | **<0.001** |
| Rural | 3708 (21.2%) | 270 (33.1%) | 3438 (20.6%) |  |
| Suburban | 3298 (18.8%) | 199 (24.4%) | 3099 (18.5%) |  |
| Urban | 10516 (60.0%) | 346 (42.5%) | 10170 (60.9%) |  |
| **Month of First Dose** |  |  |  |  |
| Dec 2020 |  |  | 2612 (15.6%) |  |
| Jan 2021 |  |  | 5358 (32.1%) |  |
| Feb 2021 |  |  | 2536 (15.2%) |  |
| Mar 2021 |  |  | 4721 (28.3%) |  |
| Apr 2021 |  |  | 1074 (6.4%) |  |
| May 2021 |  |  | 141 (0.8%) |  |
| Jun 2021 |  |  | 65 (0.4%) |  |
| Jul 2021 |  |  | 117 (0.7%) |  |
| Aug 2021 |  |  | 83 (0.5%) |  |
| **Vaccine Product** |  |  |  |  |
| Pfizer-BioNTech |  |  | 10825 (64.8%) |  |
| Moderna |  |  | 4775 (28.6%) |  |
| Janssen (Johnson & Johnson) |  |  | 746 (4.5%) |  |
| Missing |  |  | 325 (1.9%) |  |
| Other |  |  | 36 (0.2%) |  |

\*Fully or partially vaccinated; having received at least one dose of a COVID-19 vaccine.
<sup>†</sup>Participants enrolled from ten sites: Atrium Health (Charlotte, NC), Campbell University (Lillington, NC), Medstar Health (Columbia, MD), New Hanover Regional Medical Center (Wilmington, NC), Tulane University (New Orleans, LA), University of Maryland – Baltimore (Baltimore, MD), University of Mississippi (Jackson, MS), Vidant Health (Greenville, NC), Wake Forest Baptist Health (Winston-Salem, NC), WakeMed Health & Hospitals (Winston-Salem, NC). http://www.covid19communitystudy.org/.
\*Other race ethnicity includes American Indian or Alaskan Native (0.2%), Asian or Pacific Islander (1.5%), mixed race/ethnicity (1.3%), and participants who chose not to specify their race/ethnicity 0.4%.

**Figure 1:**
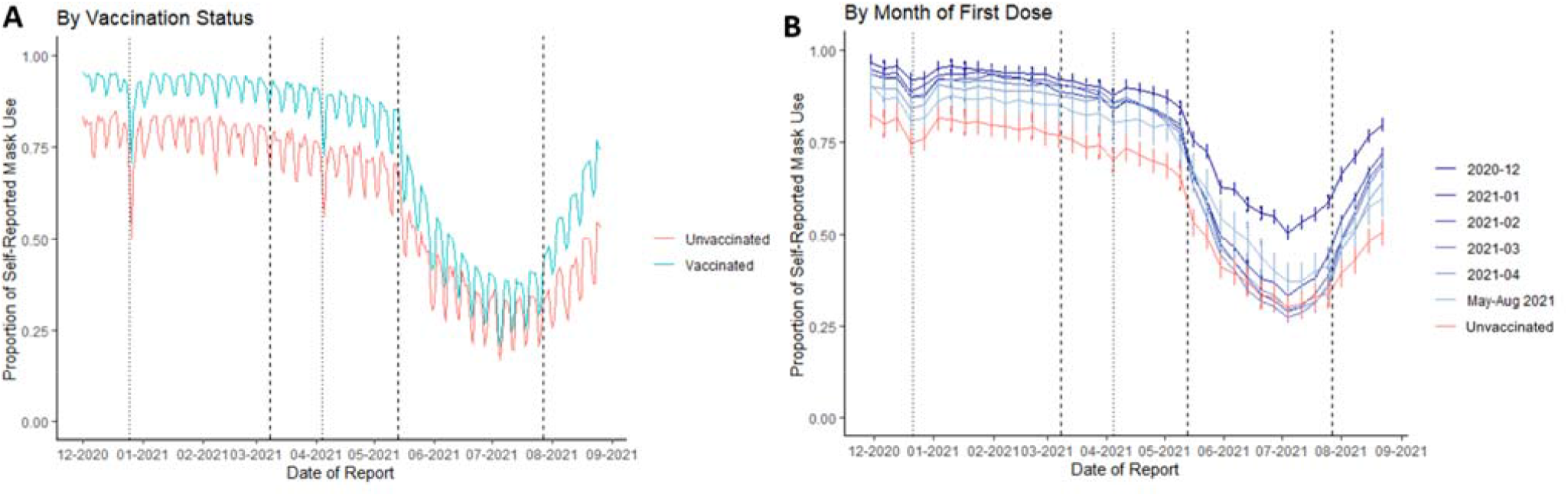
Temporal trends in the proportion of respondents self-reporting mask use. Left panel A: By vaccination status as of August 31, 2021, per day. Data shown are daily sample proportion. Right panel B: By month of first vaccine, smoothed by week. Data shown are sample proportion per week with 95% confidence intervals (calculated using standard normal approximation to the binomial distribution). Dashed reference lines drawn at March 8, 2021^7^ and May 13, 2021^4,5^, and July 27, 2021^6^, the dates corresponding to changes in CDC community masking guidance. Dotted reference lines are drawn at December 25, 2020 (Christmas Day) and April 4, 2021 (Easter Day).

Figure 1 shows mask use over time by day among vaccinated and unvaccinated participants (A) and smoothed by week, stratified by month of first vaccination (B). Mask use remained relatively stable from December 2020 through April 2021 (71-80% in unvaccinated; 86-93% in vaccinated) then declined in all groups beginning in mid-May 2021, to 34% in unvaccinated and 42% in vaccinated in June 2021, before increasing again by August (to 41% and 60% respectively). Overall, unvaccinated participants reported a mean of 12.5% less mask use across the study period, and those vaccinated in spring 2021 reported slightly less mask use compared to those vaccinated earlier. In all groups, mask use declined on weekends, holidays, and in May 2021 after the CDC recommended that fully vaccinated individuals need not wear masks in most indoor settings,^4,5^ and increased in late July 2021 after CDC recommended indoor mask use for all persons in areas of substantial or high COVID-19 transmission^6^ (Figure 1, Supplemental Table 3). The number of participants reporting “no interactions” remained relatively stable over time (Supplemental Figure 3) and a similar trend in mask use was observed when “no interactions” was combined with “yes” (Supplemental Figure 4).

Vaccination status independently predicted higher mask use after adjusting for self-reported prior COVID-19 infection, age, sex, race and ethnicity, and healthcare worker occupation during the first weeks of March, June and August, after adjusting for age, sex, race/ethnicity, and working in healthcare (Supplemental Table 1, Supplemental Table 4).

## Discussion

Mask use during non-household exposures was consistently higher among those who self-reported receiving at least one dose of COVID-19 vaccine than those who did not. Our data show changes in mask use over time, perhaps reflecting the perceived risk of infection and changes in public health messaging^8^. Although there was a previous gradual decline in mask use, the sharp decline in May coincided with updated guidance on mask use for fully vaccinated individuals^4,5^. However, we observed a simultaneous decline among unvaccinated individuals that was nearly as large, indicating that despite risk, they too relaxed mask use. The increase in mask use starting in late July, which was more pronounced in vaccinated individuals, coincides with increased transmission associated with the Delta variant and updated recommendations on mask use in indoor and public settings^5^.

This study included a large number of geographically diverse participants, and prospective data collection over nine months. However, generalization of these findings is limited by selection, reporting, and response biases, as well as the study population, which had limited numbers of participants from racial and ethnic minority groups. Participants voluntarily completed daily surveys and are affiliated with healthcare systems; therefore, they may be more engaged in healthcare than the general population. This may also explain the high rate of vaccination (95%) among participants compared to the general population (75%)^9^. Additionally, the survey question asks about mask use in “all” interactions; this does not allow for reporting varying masking behaviors in different settings and does not include a “sometimes” response which may more closely align with the behaviors of many participants.

Our data demonstrate a consistent temporal association between mask use and vaccination status. The relatively stable rate of reported interactions (Supplemental Figure 2) outside the household indicates that mask use changed independently of interactions outside the household. Although mask use declined on weekends and holidays, use remained higher among vaccinated participants. Higher mask use among those vaccinated earlier might reflect higher mask use among healthcare workers and those aged ≥65 years who became eligible for vaccination before other groups.

Our findings have several implications for public health. Mask use was lowest on weekends and holidays, reinforcing the need for messaging to maintain precautions during this time, especially during periods of high transmission. Risks of transmission are also likely to be elevated among people who are younger, male, or White, similar to previous findings^10^. Because the risks of transmission and poor outcomes are much higher if unvaccinated, the lower rate of mask use in this group is particularly concerning. We found some evidence of responsiveness to public health masking recommendations. However, our findings also highlight the need to target messaging to those least likely to wear masks and those who are unvaccinated.

In conclusion, unvaccinated participants consistently reported lower rates of mask use during non-household encounters compared with vaccinated participants. Mask use decreased during May and increased again in late July 2021, suggesting that public health guidelines affect preventive behavior. These findings underscore the need to optimize public health messaging and interventions to promote and encourage protective health behaviors and strategies amid the continued threat of emerging COVID-19 variants.

## Data Availability

The datasets used and/or analyzed during the current study are available from the corresponding author on reasonable request.

## Acknowledgements

The COVID-19 Community Research Partnership gratefully acknowledges the commitment and dedication of the study participants. Programmatic, laboratory, and technical support was provided by Vysnova Partners, Inc., Javara, Inc., and Oracle Corporation.

## Disclaimer

The findings and conclusions in this report are those of the author(s) and do not necessarily represent the official position of the Centers for Disease Control and Prevention (CDC).

## Declarations

### Ethics approval

Activity was determined to meet the definition of research [45 CFR 46.102(l)] involving human subjects [45 CFR 46.102 (e)(1)] and Institutional Review Board (IRB) approval was provided by Wake Forest University.

### Consent to participate

All participants in the COVID-19 Community Research Partnership provided written consent for participation.

### Consent for publication

Not applicable. No identifying data from any individual person is contained in the manuscript.

### Code availability

Not applicable.

Conflicts of interest/Financial disclosures: JEP owns common stock in Pfizer, Inc. The other authors declare that they have no competing interests.

### Authors’ contributions

Concept/Design: LC, AHT, SLE, MR

Data analysis/Interpretation: LC, AHT, SLE, MS Drafting article: LC, AHT, SLE, MR

Critical revision of article: All authors

Approval of article: All authors

Statistics: AHT, SLE, MS

Data collection: All authors

## Authorship Appendix

The COVID-19 Research Group (*Site Principal Investigator)

### Wake Forest School of Medicine

Thomas F Wierzba PhD, MPH, MS*, John Walton Sanders, MD, MPH, David Herrington, MD, MHS, Mark A. Espeland, PhD, MA, John Williamson, PharmD, Morgana Mongraw-Chaffin, PhD, MPH, Alain Bertoni, MD, MPH, Martha A. Alexander-Miller, PhD, Paola Castri, MD, PhD, Allison Mathews, PhD, MA, Iqra Munawar, MS, Austin Lyles Seals, MS, Brian Ostasiewski, Christine Ann Pittman Ballard, MPH, Metin Gurcan, PhD, MS, Alexander Ivanov, MD, Giselle Melendez Zapata, MD, Marlena Westcott, PhD, Karen Blinson, Laura Blinson, Mark Mistysyn, Donna Davis, Lynda Doomy, Perrin Henderson, MS, Alicia Jessup, Kimberly Lane, Beverly Levine, PhD, Jessica McCanless, MS, Sharon McDaniel, Kathryn Melius, MS, Christine O’Neill, Angelina Pack, RN, Ritu Rathee, RN, Scott Rushing, Jennifer Sheets, Sandra Soots, RN, Michele Wall, Samantha Wheeler, John White, Lisa Wilkerson, Rebekah Wilson, Kenneth Wilson, Deb Burcombe, Georgia Saylor, Megan Lunn, Karina Ordonez, Ashley O’Steen, MS, Leigh Wagner.

### Atrium Health

Michael S. Runyon MD, MPH*, Lewis H. McCurdy MD*, Michael A. Gibbs, MD, Yhenneko J. Taylor, PhD, Lydia Calamari, MD, Hazel Tapp, PhD, Amina Ahmed, MD, Michael Brennan, DDS, Lindsay Munn, PhD RN, Keerti L. Dantuluri, MD, Timothy Hetherington, MS, Lauren C. Lu, Connell Dunn, Melanie Hogg, MS, CCRA, Andrea Price, Marina Leonidas, Melinda Manning, Whitney Rossman, MS, Frank X. Gohs, MS, Anna Harris, MPH, Jennifer S. Priem, PhD, MA, Pilar Tochiki, Nicole Wellinsky, Crystal Silva, Tom Ludden PhD, Jackeline Hernandez, MD, Kennisha Spencer, Laura McAlister.

### MedStar Health Research Institute

William Weintraub MD*, Kristen Miller, DrPH, CPPS*, Chris Washington, Allison Moses, Sarahfaye Dolman, Julissa Zelaya-Portillo, John Erkus, Joseph Blumenthal, Ronald E. Romero Barrientos, Sonita Bennett, Shrenik Shah, Shrey Mathur, Christian Boxley, Paul Kolm, PhD, Ella Franklin, Naheed Ahmed, Moira Larsen.

### Tulane

Richard Oberhelman MD*, Joseph Keating PhD*, Patricia Kissinger, PhD, John Schieffelin, MD, Joshua Yukich, PhD, Andrew Beron, MPH, Johanna Teigen, MPH.

### University of Maryland School of Medicine

Karen Kotloff MD*, Wilbur H. Chen MD, MS*, DeAnna Friedman-Klabanoff, MD, Andrea A. Berry, MD, Helen Powell, PhD, Lynnee Roane, MS, RN, Reva Datar, MPH, Colleen Reilly.

### University of Mississippi

Adolfo Correa MD, PhD*, Bhagyashri Navalkele, MD, Yuan-I Min, PhD, Alexandra Castillo, MPH, Lori Ward, PhD, MS, Robert P. Santos, MD, MSCS, Pramod Anugu, Yan Gao, MPH, Jason Green, Ramona Sandlin, RHIA, Donald Moore, MS, Lemichal Drake, Dorothy Horton, RN, Kendra L. Johnson, MPH, Michael Stover.

### Wake Med Health and Hospitals

William H. Lagarde MD*, LaMonica Daniel, BSCR.

### New Hanover

Patrick D. Maguire MD*, Charin L. Hanlon, MD, Lynette McFayden, MSN, CCRP, Isaura Rigo, MD, Kelli Hines, BS, Lindsay Smith, BA, Monique Harris, CCRP, Belinda Lissor, AAS, CCRP, Vivian Cook, MA, MPH, Maddy Eversole, BS, Terry Herrin, BS, Dennis Murphy, RN, Lauren Kinney, BS, Polly Diehl, MS, RHIA, Nicholas Abromitis, BS, Tina St. Pierre, BS, Bill Heckman, Denise Evans, Julian March, BA, Ben Whitlock, CPA, MSA, Wendy Moore, BS, AAS, Sarah Arthur, MSW, LCSW, Joseph Conway.

### Vidant Health

Thomas R. Gallaher MD*, Mathew Johanson, MHA, CHFP, Sawyer Brown, MHA, Tina Dixon, MPA, Martha Reavis, Shakira Henderson, PhD, DNP, MS, MPH, Michael Zimmer, PhD, Danielle Oliver, Kasheta Jackson, DNP, RN, Monica Menon, MHA, Brandon Bishop, MHA, Rachel Roeth, MHA.

### Campbell University School of Osteopathic Medicine

Robin King-Thiele DO*, Terri S. Hamrick PhD*, Abdalla Ihmeidan, MHA, Amy Hinkelman, PhD, Chika Okafor, MD (Cape Fear Valley Medical Center), Regina B. Bray Brown, MD, Amber Brewster, MD, Danius Bouyi, DO, Katrina Lamont, MD, Kazumi Yoshinaga, DO, (Harnett Health System), Poornima Vinod, MD, A. Suman Peela, MD, Giera Denbel, MD, Jason Lo, MD, Mariam Mayet-Khan, DO, Akash Mittal, DO, Reena Motwani, MD, Mohamed Raafat, MD (Southeastern Health System), Evan Schultz, DO, Aderson Joseph, MD, Aalok Parkeh, DO, Dhara Patel, MD, Babar Afridi, DO (Cumberland County Hospital System, Cape Fear Valley).

### George Washington University Data Coordinating Center

Diane Uschner PhD*, Sharon L. Edelstein, ScM, Michele Santacatterina, PhD, Greg Strylewicz, PhD, Brian Burke, MS, Mihili Gunaratne, MPH, Meghan Turney, MA, Shirley Qin Zhou, MS, Ashley H Tjaden, MPH, Lida Fette, MS, Asare Buahin, Matthew Bott, Sophia Graziani, Ashvi Soni, MS.

### George Washington University Mores Lab

Christopher Mores, PhD, Abigail Porzucek, MS.

### Oracle Corporation

Rebecca Laborde, Pranav Acharya.

### Sneez LLC

Lucy Guill, MBA, Danielle Lamphier, MBA, Anna Schaefer, MSM, William M. Satterwhite, JD, MD.

### Vysnova Partners

Anne McKeague, PhD, Johnathan Ward, MS, Diana P. Naranjo, MA, Nana Darko, MPH, Kimberly Castellon, BS, Ryan Brink, MSCM, Haris Shehzad, MS, Derek Kuprianov, Douglas McGlasson, MBA, Devin Hayes, BS, Sierra Edwards, MS, Stephane Daphnis, MBA, Britnee Todd, BS.

### Javara Inc

Atira Goodwin.

### External Advisory Council

Ruth Berkelman, MD, Emory, Kimberly Hanson, MD, U of Utah, Scott Zeger, PhD, Johns Hopkins, Cavan Reilly, PhD, U. of Minnesota, Kathy Edwards, MD, Vanderbilt, Helene Gayle, MD MPH, Chicago Community Trust, Stephen Redd.

## Masking Behavior: Supplemental Tables and Figures

**Supplemental Table 1:** COVID-19 Community Research Partnership Multivariable logistic regression models predicting “Yes” response to mask use question at least one day during select weeks in 2021

|  | Odds Ratio | Lower<br>95% CI | Upper<br>95% CI | p-value |
| --- | --- | --- | --- | --- |
| **First week of March 2021:** |  |  |  |  |
| **Vaccinated by March 1, 2021** (Yes vs No) | 3.58 | 2.73 | 4.68 | <0.001 |
| **Self-reported positive COVID-19 test prior to March 1** (Yes vs No) | 0.96 | 0.69 | 1.34 | 0.81 |
| **Age Group** (35-44 vs 18-34 years) | 1.34 | 0.94 | 1.91 | 0.10 |
| **Age Group** (45-64 vs 18-34 years) | 1.43 | 1.05 | 1.95 | 0.02 |
| **Age Group** (65+ vs 18-34 years) | 1.61 | 1.15 | 2.26 | 0.01 |
| **Sex** (Male vs Female) | 1.05 | 0.87 | 1.28 | 0.61 |
| **Race/Ethnicity** (Hispanic vs NH White) | 0.72 | 0.41 | 1.27 | 0.25 |
| **Race/ Ethnicity** (NH Black vs NH White) | 1.08 | 0.67 | 1.72 | 0.76 |
| **Race/ Ethnicity** (Other vs NH White) | 0.81 | 0.51 | 1.28 | 0.36 |
| **Healthcare Worker** (Yes vs No) | 2.29 | 1.77 | 2.97 | <0.001 |
| **First week of June 2021:** |  |  |  |  |
| **Vaccinated by June 1, 2021** (Yes vs No) | 1.86 | 1.59 | 2.18 | <0.001 |
| **Self-reported positive COVID-19 test prior to June 1** (Yes vs No) | 0.67 | 0.58 | 0.77 | <0.001 |
| **Age Group** (35-44 vs 18-34 years) | 0.94 | 0.79 | 1.11 | 0.46 |
| **Age Group** (45-64 vs 18-34 years) | 0.94 | 0.81 | 1.10 | 0.44 |
| **Age Group** (65+ vs 18-34 years) | 1.19 | 1.01 | 1.40 | 0.04 |
| **Sex** (Male vs Female) | 0.74 | 0.68 | 0.80 | <0.001 |
| **Race/Ethnicity** (Hispanic vs NH White) | 1.39 | 1.02 | 1.90 | 0.04 |
| **Race/ Ethnicity** (NH Black vs NH White) | 2.91 | 2.20 | 3.86 | <0.001 |
| **Race/ Ethnicity** (Other vs NH White) | 1.47 | 1.15 | 1.86 | <0.001 |
| **Healthcare Worker** (Yes vs No) | 1.65 | 1.49 | 1.82 | <0.001 |
| **First week of August 2021:** |  |  |  |  |
| **Vaccinated by August 1, 2021** (Yes vs No) | 1.93 | 1.65 | 2.25 | <0.001 |
| **Self-reported positive COVID-19 test prior to August 1** (Yes vs No) | 0.65 | 0.57 | 0.74 | <0.001 |
| **Age Group** (35-44 vs 18-34 years) | 1.02 | 0.87 | 1.19 | 0.81 |
| **Age Group** (45-64 vs 18-34 years) | 1.10 | 0.96 | 1.27 | 0.18 |
| **Age Group** (65+ vs 18-34 years) | 1.62 | 1.39 | 1.89 | <0.001 |
| **Sex** (Male vs Female) | 0.72 | 0.66 | 0.78 | <0.001 |
| **Race/Ethnicity** (Hispanic vs NH White) | 1.83 | 1.32 | 2.53 | <0.001 |
| **Race/ Ethnicity** (NH Black vs NH White) | 2.85 | 2.18 | 3.72 | <0.001 |
| **Race/ Ethnicity** (Other vs NH White) | 1.44 | 1.15 | 1.81 | <0.001 |
| **Healthcare Worker** (Yes vs No) | 1.52 | 1.38 | 1.67 | <0.001 |

Multivariable logistic regression modelling any mask use during selected weeks adjusted for vaccination status and self-reported positive test prior to date of interest, age group, sex, race/ethnicity, and healthcare worker occupation.

**Supplemental Table 2:** Comparison of Characteristics for Participants Included in Current Analysis versus Other Participants Enrolled by December 2020

|  | **Not Included**<br>**(N=18698)** | **Included**<br>**(N=17522)** | **p-value** |
| --- | --- | --- | --- |
| **Age (years)** |  |  |  |
| Mean (SD) | 43.8 (15.4) | 55.3 (13.8) | <0.001 |
| **Sex** |  |  | 0.014 |
| Missing | 9 (0.0%) | 0 (0%) |  |
| Female | 12759 (68.2%) | 11932 (68.1%) |  |
| Male | 5930 (31.7%) | 5590 (31.9%) |  |
| **Race/Ethnicity** |  |  | <0.001 |
| Hispanic, Any Race | 552 (3.0%) | 326 (1.9%) |  |
| Non-Hispanic Black | 1302 (7.0%) | 728 (4.2%) |  |
| Non-Hispanic White | 15782 (84.4%) | 15884 (90.7%) |  |
| Other | 1062 (5.7%) | 584 (3.3%) |  |
| **Health Care Worker** |  |  | <0.001 |
| No | 13464 (72.0%) | 13036 (74.4%) |  |
| Yes | 5234 (28.0%) | 4486 (25.6%) |  |
| **County Classification** |  |  | <0.001 |
| Missing | 30 (0.2%) | 0 (0%) |  |
| Rural | 3917 (20.9%) | 3708 (21.2%) |  |
| Suburban | 3566 (19.1%) | 3298 (18.8%) |  |
| Urban | 11185 (59.8%) | 10516 (60.0%) |  |

**Supplemental Table 3.** Mask Use Comparison at Select Times

| **Time Frames** | **Overall Mask Use** |
| --- | --- |
| **Christmas Holiday** |  |
| December 25, 2020 | 69.4% |
| December 1-24, 2020 | 92.6% |
| **Easter Holiday** |  |
| April 4, 2021 | 72.1% |
| March 14-April 3, 2021 | 87.9% |
| **Weekend/Weekday** |  |
| Weekend | 66.4% |
| Weekday | 75.0% |
| **CDC Guidelines May 13, 2021** |  |
| April 12 – May 12, 2021 | 83.9% |
| May 14 – June 14, 2021 | 53.4% |

**Supplemental Table 4.** COVID-19 Community Research Partnership Multivariable logistic regression models predicting “Yes” or “No Interactions” responses to mask use question at least one day during select weeks in 2021.

|  | **Odds Ratio** | **Lower 95% CI** | **Upper 95% CI** | **p-value** |
| --- | --- | --- | --- | --- |
| **First week of March 2021:** |  |  |  |  |
| **Vaccinated by March 1, 2021** (Yes vs No) | 9.02 | 6.35 | 12.82 | <0.001 |
| **Self-reported positive COVID-19 test prior to March 1** (Yes vs No) | 0.93 | 0.57 | 1.51 | 0.76 |
| **Age Group** (35-44 vs 18-34 years) | 1.23 | 0.71 | 2.13 | 0.46 |
| **Age Group** (45-64 vs 18-34 years) | 1.47 | 0.90 | 2.41 | 0.13 |
| **Age Group** (65+ vs 18-34 years) | 2.68 | 1.46 | 4.90 | <0.001 |
| **Sex** (Male vs Female) | 0.61 | 0.44 | 0.84 | <0.001 |
| **Race/Ethnicity** (Hispanic vs NH White) | 0.85 | 0.31 | 2.35 | 0.76 |
| **Race/ Ethnicity** (NH Black vs NH White) | 1.48 | 0.60 | 3.64 | 0.40 |
| **Race/ Ethnicity** (Other vs NH White) | 1.08 | 0.47 | 2.48 | 0.86 |
| **Healthcare Worker** (Yes vs No) | 1.19 | 0.82 | 1.72 | 0.36 |
| **First week of June 2021:** |  |  |  |  |
| **Vaccinated by June 1, 2021** (Yes vs No) | 1.72 | 1.46 | 2.03 | <0.001 |
| **Self-reported positive COVID-19 test prior to June 1** (Yes vs No) | 0.64 | 0.56 | 0.74 | <0.001 |
| **Age Group** (35-44 vs 18-34 years) | 0.92 | 0.77 | 1.10 | 0.35 |
| **Age Group** (45-64 vs 18-34 years) | 0.91 | 0.78 | 1.07 | 0.26 |
| **Age Group** (65+ vs 18-34 years) | 1.19 | 1.00 | 1.41 | 0.05 |
| **Sex** (Male vs Female) | 0.71 | 0.65 | 0.77 | <0.001 |
| **Race/Ethnicity** (Hispanic vs NH White) | 1.57 | 1.13 | 2.19 | 0.01 |
| **Race/ Ethnicity** (NH Black vs NH White) | 3.66 | 2.65 | 5.06 | <0.001 |
| **Race/ Ethnicity** (Other vs NH White) | 1.86 | 1.42 | 2.44 | <0.001 |
| **Healthcare Worker** (Yes vs No) | 1.60 | 1.44 | 1.77 | <0.001 |
| **First week of August 2021:** |  |  |  |  |
| **Vaccinated by August 1, 2021** (Yes vs No) | 1.86 | 1.59 | 2.18 | <0.001 |
| **Self-reported positive COVID-19 test prior to August 1** (Yes vs No) | 0.65 | 0.56 | 0.74 | <0.001 |
| **Age Group** (35-44 vs 18-34 years) | 0.98 | 0.83 | 1.16 | 0.83 |
| **Age Group** (45-64 vs 18-34 years) | 1.08 | 0.94 | 1.26 | 0.28 |
| **Age Group** (65+ vs 18-34 years) | 1.59 | 1.35 | 1.86 | <0.001 |
| **Sex** (Male vs Female) | 0.68 | 0.62 | 0.74 | <0.001 |
| **Race/Ethnicity** (Hispanic vs NH White) | 1.98 | 1.41 | 2.79 | <0.001 |
| **Race/ Ethnicity** (NH Black vs NH White) | 4.58 | 3.26 | 6.42 | <0.001 |
| **Race/ Ethnicity** (Other vs NH White) | 1.68 | 1.32 | 2.16 | <0.001 |
| **Healthcare Worker** (Yes vs No) | 1.45 | 1.32 | 1.60 | <0.001 |

Multivariable logistic regression modelling any mask use (“Yes” and “No interactions”) during selected weeks adjusted for vaccination status and self-reported positive test prior to date of interest, age group, sex, race/ethnicity, and healthcare worker occupation.

**Supplemental Figure 1.**
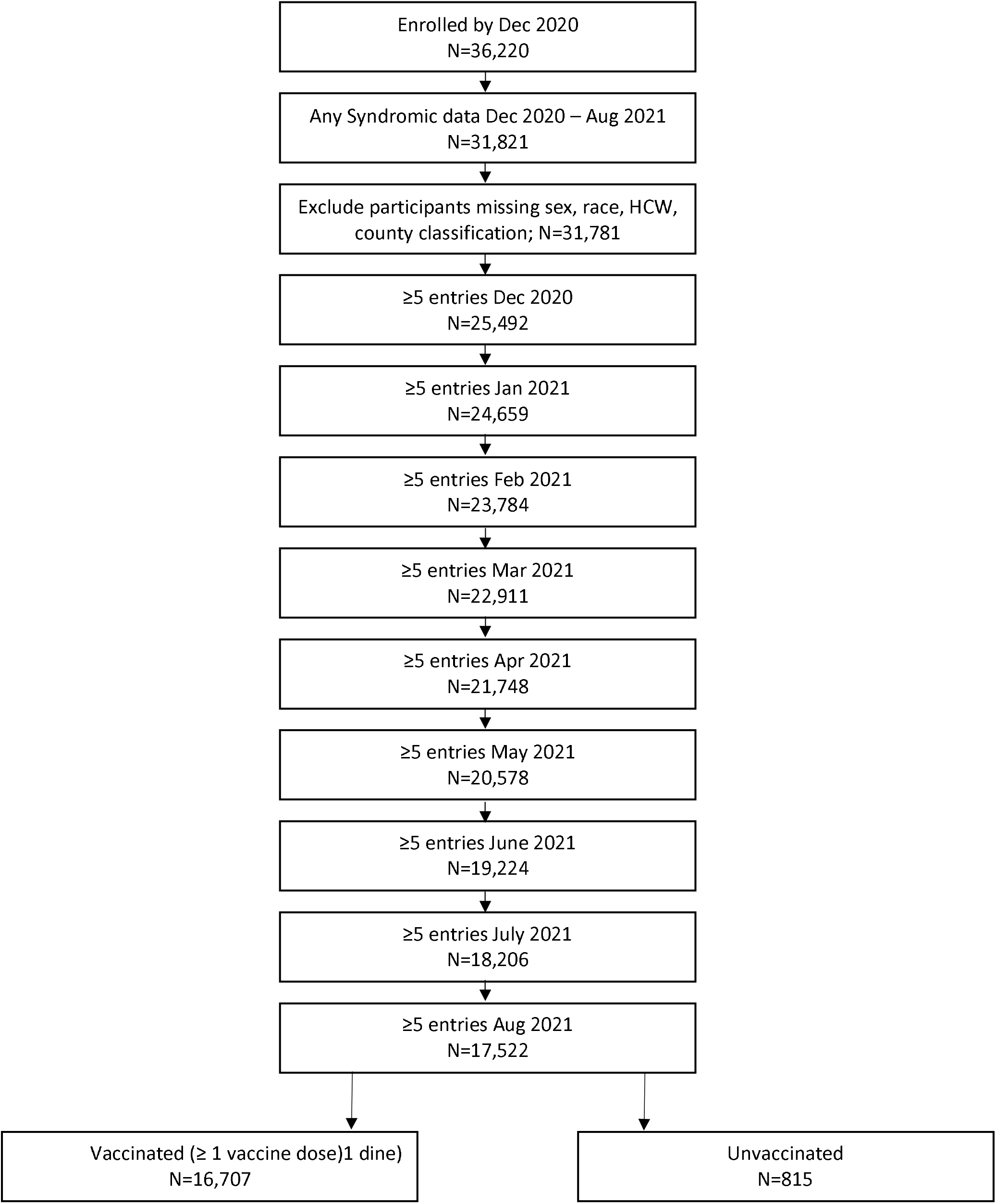
Inclusion Flow Diagram

**Supplemental Figure 2:**
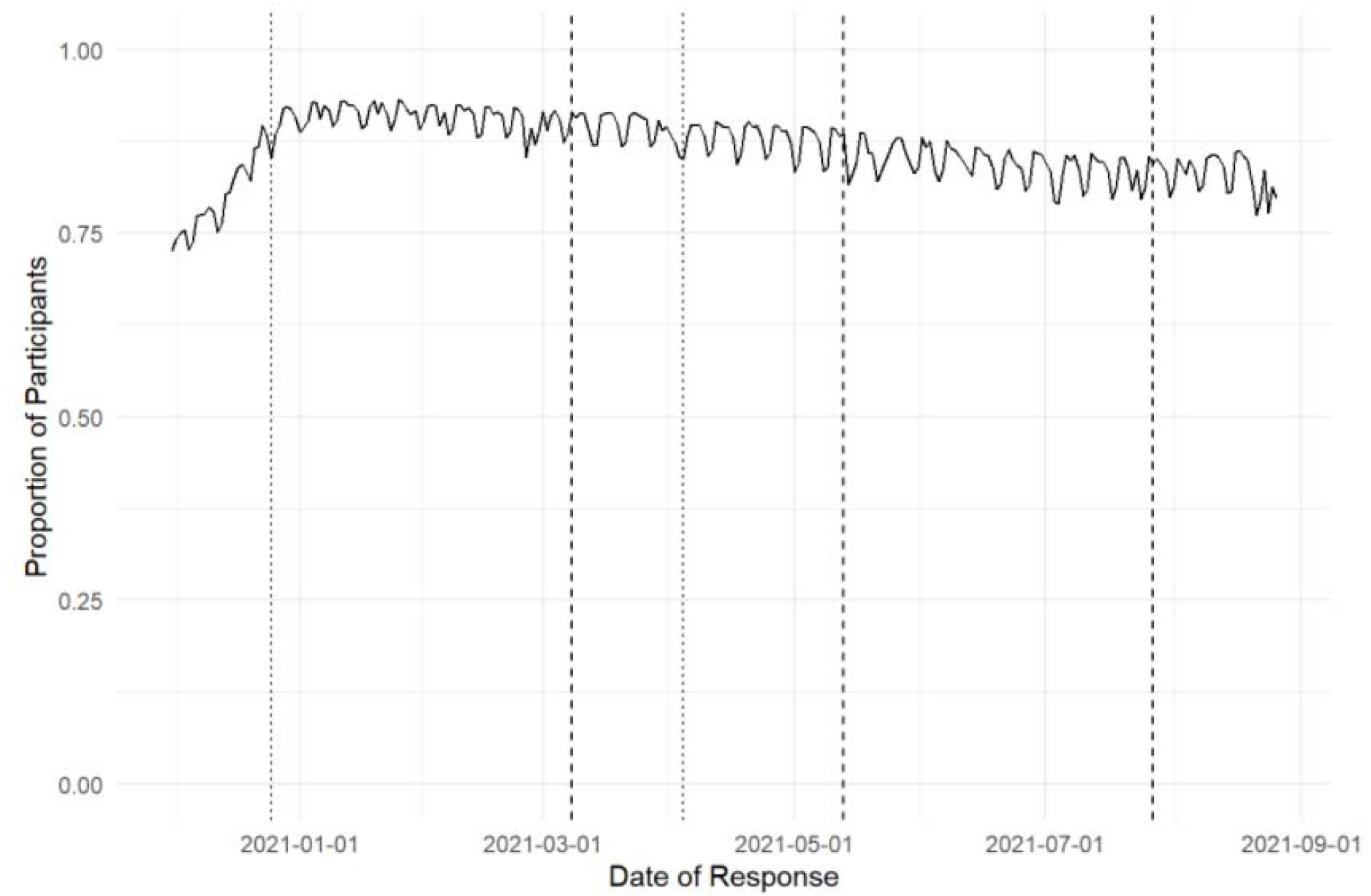
Responses to daily survey over time. Shown is the proportion of participants who responded to the daily survey among the total analytic sample. Dashed reference lines are drawn at March 8, 2021^5^ and May 13, 2021^2,3^, and July 27, 2021^4^, the dates corresponding to changes in CDC community masking guidance. Dotted reference lines are drawn at December 25, 2020 (Christmas Day) and April 4, 2021 (Easter Day).

**Supplemental Figure 3.**
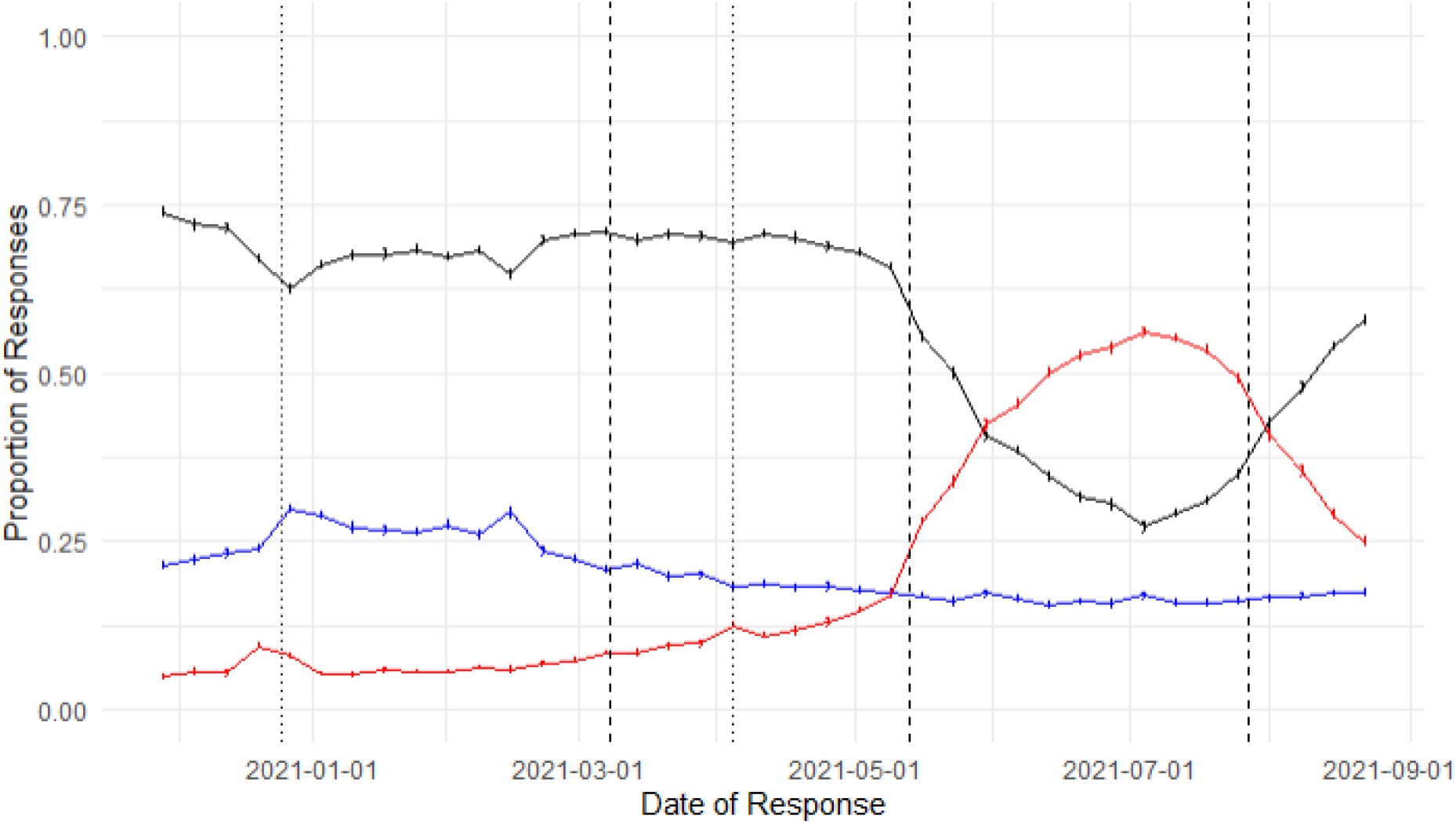
Overall proportion of respondents self-reporting mask use, no interactions, and no mask use over time. Black=Yes; Blue=No Interactions; Red=No. Data shown are weekly mean proportion of responses with 95% CI. Dashed reference lines drawn at March 8, 2021^5^ and May 13, 2021^2,3^, and July 27, 2021^4^, the dates corresponding to changes in CDC community masking guidance. Dotted reference lines are drawn at December 25, 2020 (Christmas Day) and April 4, 2021 (Easter Day).

**Supplemental Figure 4:**
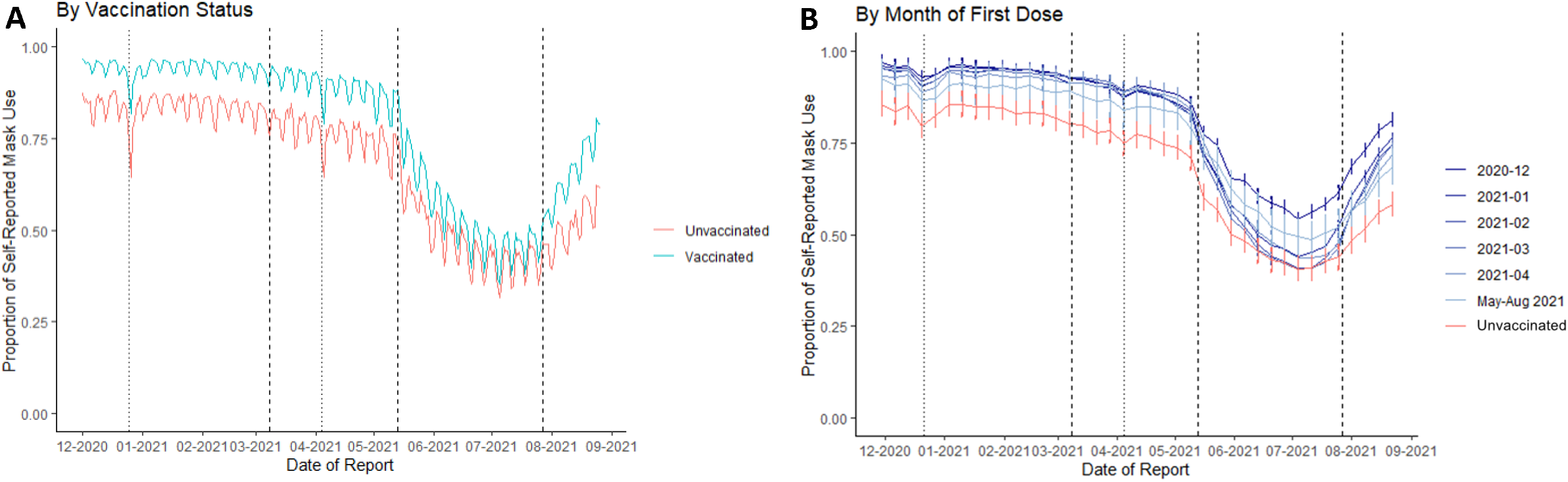
Temporal trends in the proportion of respondents self-reporting mask use (Responding “Yes” or “No Interactions”). Left panel A: By vaccination status as of August 31, 2021, per day. Data shown are daily sample proportion. Right panel B: By month of first vaccine, smoothed by week. Data shown are weekly sample proportion with 95% CI (calculated using standard normal approximation to the binomial distribution). Dashed reference lines drawn at March 8, 2021^5^ and May 13, 2021^2,3^, and July 27, 2021^4^, the dates corresponding to changes in CDC community masking guidance. Dotted reference lines are drawn at December 25, 2020 (Christmas Day) and April 4, 2021 (Easter Day)

